# Depression and Anxiety Symptoms in Young Adults Before and During the COVID-19 Pandemic: Evidence from a Canadian Population-Based Cohort

**DOI:** 10.1101/2021.04.23.21255994

**Authors:** Kia Watkins-Martin, Massimiliano Orri, Marie-Hélène Pennestri, Natalie Castellanos-Ryan, Simon Larose, Jean-Philippe Gouin, Isabelle Ouellet-Morin, Nicholas Chadi, Frederick Philippe, Michel Boivin, Richard E. Tremblay, Sylvana Côté, Marie-Claude Geoffroy

## Abstract

**Objectives:** Concerns have been raised that the COVID-19 pandemic could increase risk for adverse mental health outcomes, especially in young adults, a vulnerable age group. We investigated changes in depression and anxiety symptoms (overall and severe) from before to during the pandemic, as well as whether these changes are linked to COVID-19 related stressors and pre-existing vulnerabilities in young adults followed in the context of a population-based cohort.

**Method:** Participants (n=1039) from the Quebec Longitudinal Study of Child Development reported on their depression and anxiety symptoms and completed a COVID-19 questionnaire during the first wave of the COVID-19 pandemic in the summer of 2020 (age 22 years). Assessments at age 20 (2018) were used to estimate pre-pandemic depression and anxiety symptom severity.

**Results:** While overall levels of depression and anxiety symptoms did not change, there was an increase in rates of severe depression (but not severe anxiety) from before (6.1%) to during (8.2%) the pandemic. Depressive and anxiety symptoms increased from before to during the COVID-19 pandemic among young adults with the lowest levels of symptoms before the pandemic, while they decreased among those with the highest levels of pre-existing symptoms. Youth who were living alone experienced an increase in depressive symptoms. Other COVID-19 related variables (e.g., loss of education/occupation, frequent news-seeking) and pre-existing vulnerabilities (e.g., low SES, low social support) were not associated with changes in depression or anxiety symptoms.

**Conclusion:** Depression and anxiety symptoms in young adults from Québec in Summer 2020 were comparable to symptoms reported in 2018. Most COVID-19 related stressors and pre-existing vulnerabilities were not associated with changes in symptoms. However, the increased rate of severe depression and the increase in depression and anxiety symptoms among young adults with the least mental health symptoms before the COVID-19 pandemic are concerning.

## Introduction

There is concern that the coronavirus disease 2019 (COVID-19) pandemic has negatively impacted depressive and anxiety symptoms (Amsalem et al., 2021; Holmes et al., 2020; Pfefferbaum and North, 2020), especially among young adults aged 18-25 years (Fancourt and Steptoe, 2020; Lai et al., 2020; McGinty et al., 2020; Qiu et al., 2020). While young adulthood is generally a period of good physical health, mental health problems are common (Jurewicz, 2015; Mojtabai et al., 2016). To date, most studies that have documented putative mental health consequences of the COVID-19 pandemic have relied on cross-sectional investigations of convenience samples (Lesser and Nienhuis, 2020; Mazza et al., 2020; Pierce et al., 2020; Rapisarda et al., 2020), from which it is not possible to draw conclusions about whether and how mental health has changed from pre-pandemic levels. Only longitudinal studies with pre-pandemic assessments of depression and anxiety symptoms can allow one to quantify such changes according to a clear temporal sequence and account for such sources of bias as seasonality. The few longitudinal studies with pre-pandemic assessments of depression and anxiety symptoms in young adults have yielded inconsistent findings, with some showing deterioration (Copeland et al., 2021; Shanahan et al., 2020) and others reporting no symptom change during the COVID-19 pandemic.

To our knowledge, no population-based study in the Canadian province of Quebec has examined whether the mental health of young adults has changed from before to during the first wave of the pandemic and identified factors associated with these changes. Using a longitudinal population-based cohort of young adults from Quebec, where strict lockdown measures (e.g., stay-at-home orders; social distancing; school and business closures) where implemented during the first wave of the COVID-19 pandemic, we aimed to: 1) examine whether and how depression and anxiety symptoms and the severity of these symptoms changed from the pre-pandemic (participant age: 20 years) to the intra-pandemic period during the summer of 2020 (participant age: 22 years); and 2) investigate whether these changes were linked to COVID-19 related stressors (e.g. loss of employment/education; COVID-19 related news seeking) and/or pre-existing vulnerabilities (low SES; severe depression and anxiety).

## Method

### Participants

The Québec Longitudinal Study of Child Development (QLSCD) (Orri et al., 2020) is an ongoing population-based cohort that includes 2120 participants born in 1997/98 in the province of Quebec, Canada. From July to August 2020, participants completed an online survey at age 22 years about their well-being during the Covid-19 pandemic. Of the 1593 individuals contacted, 1182 responded in 2020 (participation rate: 74%). Of those, 1039 had provided information on their mental health before the pandemic in the Spring of 2018, at age 20 years. The QLSCD, conducted by the Institut de la Statistique du Québec (ISQ; 17), was approved by ethical committees of the ISQ and the CHU Sainte-Justine Hospital Research Centre and written informed consent was obtained.

### Measures

#### Depression and anxiety symptoms before (20 years) and during (22 years) the Covid-19 pandemic

Depressive symptoms were self-reported using the Centre for Epidemiological Studies Depression Scale, short form, (CES-D) including 12 items; (e.g., “I felt depressed”) rated from 0=rarely/never to 3=most/all of the time (Ferro et al., 2015; Poulin et al., 2005; Radloff, 1977). Anxiety symptoms were self-reported using the Generalized Anxiety Disorder 7-item scale (GAD-7) including 7 items (e.g., “Feeling nervous, anxious or on edge”) rated from 0=not at all to 3=nearly every day (Spitzer et al., 2006). Scores of 21 to 36 and of 15 to 21 are thought to indicate severe levels of depression and anxiety, respectively (Radloff, 1977).

#### COVID-19 related stressors

All participants completed a questionnaire assessing their worries about the pandemic. Participants indicated their level of concern on a 4-point Likert scale (1=not at all to 4=extremely concerned) regarding: having a degree/certificate/diploma which will not be considered equivalent, compared to those who completed their degree prior to the pandemic; not having job prospects in the near future; and not having enough money to meet basic needs. The questionnaire also included items about their living status (alone vs. with others) and the level of disruption that the pandemic had on their life from mid-March to Summer 2020, including: loss of employment (“I lost my job”; “I closed my business”); loss of education (“All or some of my courses have been rescheduled to Fall 2020”; “I have dropped all my courses”; “My internship has been postponed or cancelled”); a positive COVID-19 test result (yes vs. no); Covid-19 related daily news seeking on traditional or social media (*<*2 hours vs. ≥ 2 hours per day); and participants’ geographic region based on their postal code (Montreal vs. other regions, as Montreal was the hardest hit region in the Province of Québec in March-August 2020, as per the number of confirmed cases and Covid-19 related deaths) (“Données COVID-19 par région sociosanitaire,” 2021).

#### Pre-existing vulnerabilities

Pre-existing vulnerabilities increasing risk for poor mental health (Fergusson et al., 2015; Goldman-Mellor et al., 2016; Lucassen et al., 2017; Pascoe et al., 2016; Scardera et al., 2020; Wilson et al., 2009) were assessed before the Québec government enforced lockdown measures as of mid-March 2020: not in education or employment at 22 years (just prior to the onset of the pandemic); having parents of low socioeconomic status (SES) (defined as scores ≤1 standard deviation (SD) on SES scale aggregating annual gross income, parental education level, and parental occupational prestige from ages 15 and 17 years) (Willms and Shields, 1996); sexual orientation at 17 years (same sex/bisexual/asexual vs. opposite sex; if missing at 17 years, orientation at 15 years was used); low social support at 19 years (defined as score ≤1 SD on the 10-item Social Provision Scale) (Caron, 2013); low life satisfaction at 19 years old (defined as a score of ≤5 on the following item: “Using a scale of 0 to 10, where 0 means ‘very dissatisfied’ and 10 means ‘very satisfied’, how do you feel about your life as a whole right now?”) (Thanh Tu and Desrosiers, 2019); and a chronic learning disability diagnosis as reported by the mother at 15 or 17 years (no vs. yes).

### Statistical analyses

All statistical analyses were conducted in IBM SPSS, version 26, using cohort weights to ensure representativeness of the sample. First, we described the COVID-19 related stressors and pre-existing vulnerabilities variables using counts and percentages. Second, we tested whether (a) a mean symptom score and (b) a severity category of depressive and anxiety symptoms changed from before to during the COVID-19 pandemic using paired t-tests. Third, we calculated change in depression and anxiety symptoms by subtracting the mean symptom score before the pandemic (20 years) to the mean score during the pandemic (22 years). We standardized the mean change score (change/SD of change) to ease interpretation; positive scores indicate an increase in depression and anxiety symptoms. Then, we examined crude associations between COVID-19 related stressors and pre-existing vulnerabilities variables and change in depression and anxiety symptoms, using t-tests and ANOVAs. The double-sided p-value for significance was set at .05. We used multiple imputation (MI) to generate 100 datasets to handle missing values on risk factors. Missing values ranged from 0.9% (loss of employment) to 17.9% (sexual minority); there was no missing data for COVID-19 related stressors.

## Results

Descriptive statistics on COVID-19 related stressors and pre-existing vulnerabilities are shown in **Table 1. Figure 1** depicts descriptive statistics for COVID-19-related concerns. The vast majority of young adults reported that they were “not at all concerned” about (a) having a compromised diploma, (b) loss of job prospects or (c) having enough money to meet basic needs, whereas 10.2%, 21.4% and 14.0% reported that they were ‘very or extremely concerned’, respectively. As shown in **Table 2**, there was no difference in the means of depression and anxiety symptoms from before to during the pandemic, as reported by participants in July-August 2020. However, while the prevalence of severe anxiety did not increase significantly, that of severe depression did, with 6.2% of participants reporting severe depressive symptoms before the pandemic compared to 8.1% during the pandemic (increase of 1.9%; *p*=.041).

**Table 1.** Descriptive statistics of the COVID-19 related stressors and pre-existing vulnerabilities, weighted.

| **COVID-19 Related Stressors** | *n* | % |
| --- | --- | --- |
| Sex |  |  |
| *Male* | 417 | 40.1 |
| *Female* | 622 | 59.9 |
| Living Status |  |  |
| *Alone* | 90 | 8.7 |
| *With Others* | 949 | 91.3 |
| Loss of Employment |  |  |
| *No* | 790 | 76.0 |
| *Yes* | 249 | 24.0 |
| Loss of Education |  |  |
| *No* | 805 | 77.5 |
| *Yes* | 234 | 22.5 |
| Positive COVID-19 Test |  |  |
| *No* | 1030 | 99.0 |
| *Yes* | 9 | 1.0 |
| Daily COVID-19 Related News Seeking |  |  |
| *<2 hours* | 783 | 75.0 |
| *≥2 hours* | 256 | 25.0 |
| Living in Montreal |  |  |
| *No* | 831 | 80.0 |
| *Yes* | 208 | 20.0 |
| **Pre-Existing Vulnerabilities** | *n* | % |
| Not in Education or Employed (22 years, pre-pandemic) |  |  |
| *No* | 941 | 90.6 |
| *Yes* | 98 | 9.4 |
| Low SES Families (15, 17 years)<sup>a</sup> |  |  |
| *No* | 873 | 84.0 |
| *Yes* | 166 | 16.0 |
| Sexual Minority (15, 17 years) |  |  |
| *No* | 921 | 88.6 |
| *Yes (Gay, Lesbian, Asexual)* | 118 | 11.4 |
| Low Social Support (19 years)<sup>a</sup> |  |  |
| *No* | 892 | 87.3 |
| *Yes* | 147 | 12.7 |
| Low Life Satisfaction (19 years)<sup>b</sup> |  |  |
| *No* | 901 | 86.7 |
| *Yes* | 138 | 13.3 |
| Learning Disability Diagnosis (15, 17 years) |  |  |
| *No* | 963 | 92.7 |
| *Yes* | 76 | 7.3 |
| Pre-Existing Depression Symptoms (20 years) |  |  |
| *No/low/moderate* | 975 | 93.8 |
| *Severe*<sup>c</sup> | 64 | 6.2 |
| Pre-Existing Anxiety Symptoms (20 years) |  |  |
| *No/low/moderate* | 990 | 95.3 |
| *Severe*<sup>c</sup> | 49 | 4.7 |

*Note:* Data were compiled from the final master file of the Québec Longitudinal Study of Child Development (1998–2020), Québec Government, Québec Statistic Institute.
Maximum N available ranges from 869 to 1039.
<sup>a</sup> Defined as scores <1 standard deviation of the sample mean.
<sup>b</sup> Defined as scores of 0 to 5 a scale of 0 to 10, where 0 means ‘very dissatisfied’ and 10 means ‘very satisfied’.
<sup>c</sup> Severe symptoms were defined by Centre for Epidemiological Studies-Depression scale scores ≥ 21 and Generalized Anxiety Disorder 7-item scale scores ≥ 15.

**Table 2.**
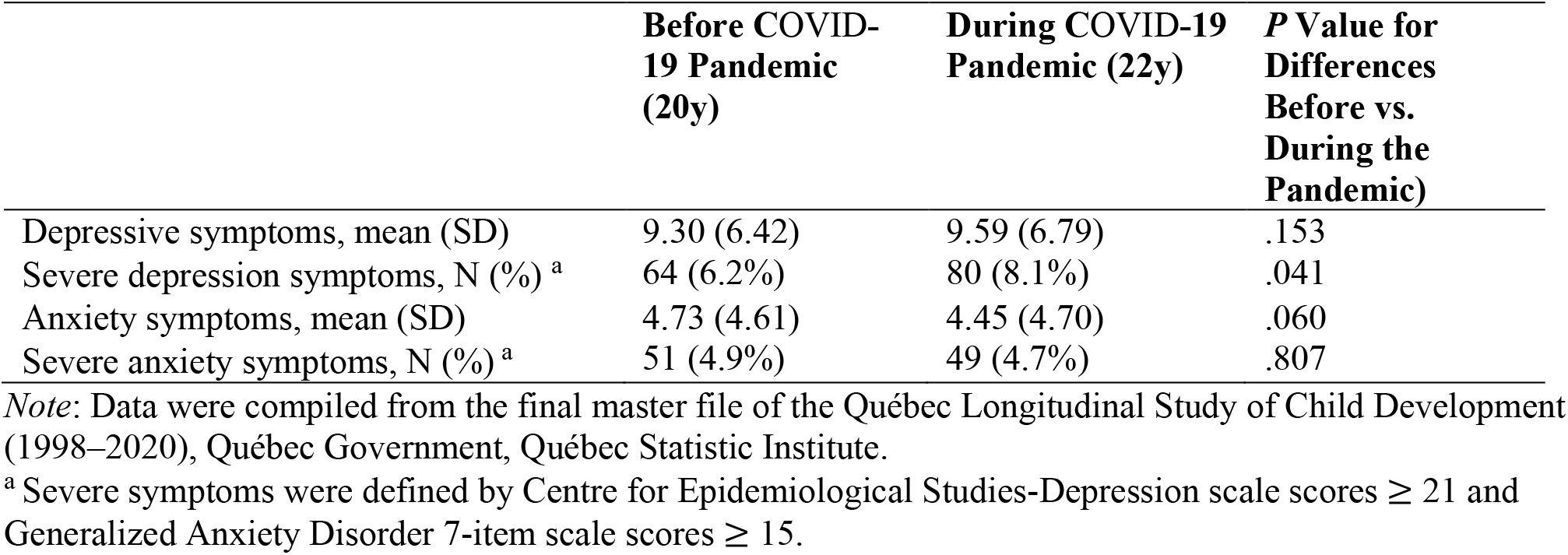
Descriptive statistics for depression and anxiety symptoms before (20 years) and during (22 years) the COVID-19 pandemic, *n*=1039, weighted.

**Figure 1.**
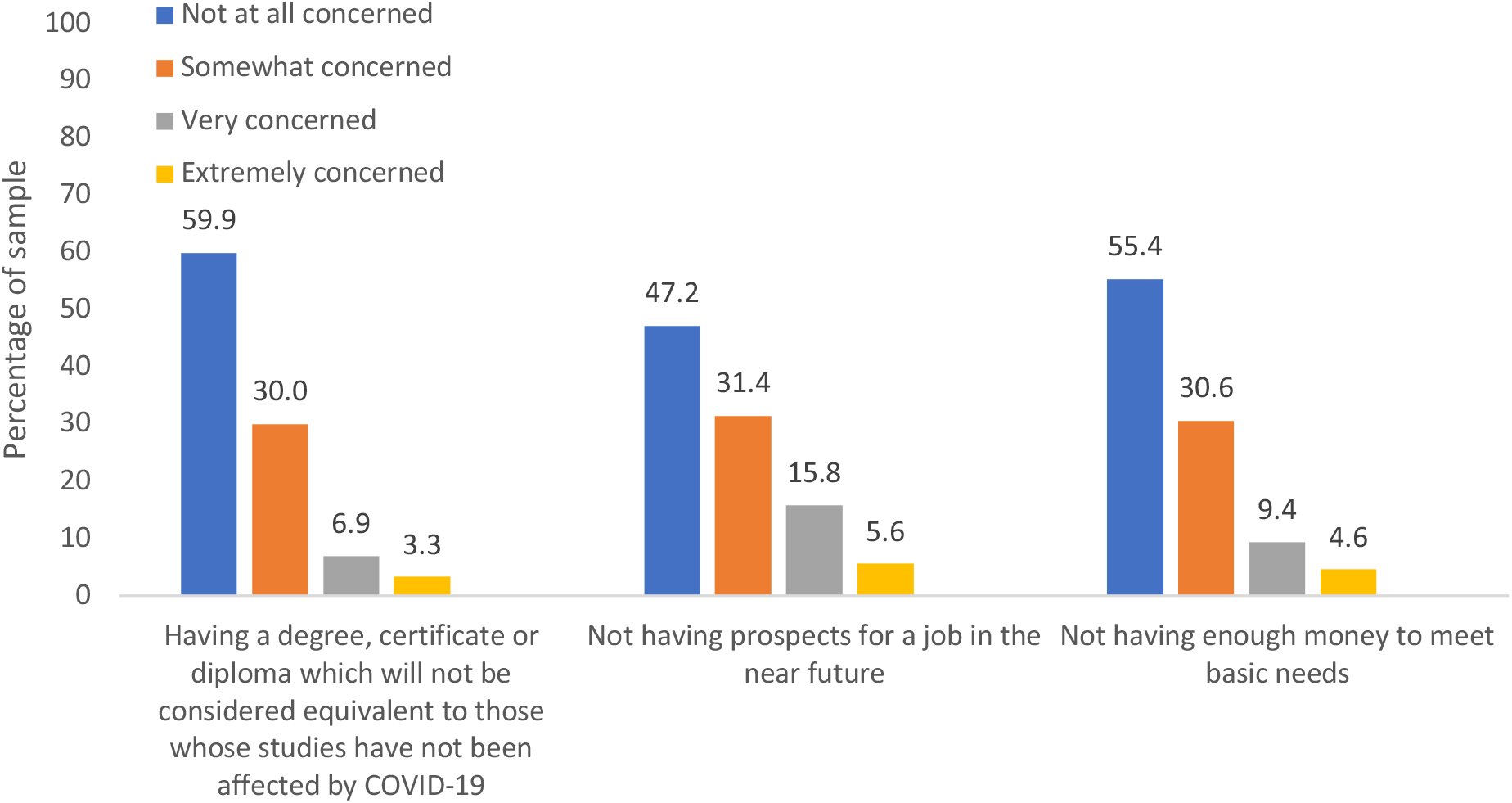
Descriptive statistics of the COVID-19-related concerns, *n*=1039, weighted *Note*: Data were compiled from the final master file of the Québec Longitudinal Study of Child Development (1998–2020), Québec Government, Québec Statistic Institute.

**Figure 2** and **Supplemental Table 1** depict standardized change in depression and anxiety from before the pandemic (20 years) to during the pandemic (22 years) as a function of Covid-related stressors. Symptoms of depression and anxiety did not vary according to COVID-19 related stressors, except for participants living alone who reported a slight increase in depressive symptoms (0.22, SD). **Figure 3** and **Supplemental Table 1** depict standardized changes in depression and anxiety symptoms as a function of pre-existing vulnerabilities. Participants with pre-existing severe symptoms of depression and anxiety experienced a significant decrease in depressive and anxiety symptoms from before to during the pandemic. That is, individuals with severe depression at age 20 years experienced a decrease in depressive symptoms from 20 to 22 years (1.26 SD), while those with severe anxiety experienced a decrease in anxiety symptoms of 1.61 SD. In sensitivity analyses, we divided participants into quintiles according to their depressive and anxiety levels at 20 years: (1) very low (0-20%), (2) low (21-40%) (3) average (41-60%), (4) high (61-80%) and (5) very high (81-100%) and explored changes in symptom levels from before to during COVID-19 using ANOVAs. As depicted in Figure 4 and Supplemental Table 2, there was a significant increase in depression and anxiety symptoms among young adults with the lowest levels of symptoms at baseline. To illustrate, participants with very low levels of depressive symptoms at age 20 years experienced a significant increase in symptoms from age 20 to 22 years (0.49 SD, *p*<.001), while those with very low levels of anxiety experienced an increase in symptoms of 0.38 SD (*p*<.001). As previously reported, young adults with the highest levels of depression and anxiety symptoms at baseline experienced a decrease in symptoms from before to during COVID-19.

**Figure 2.**
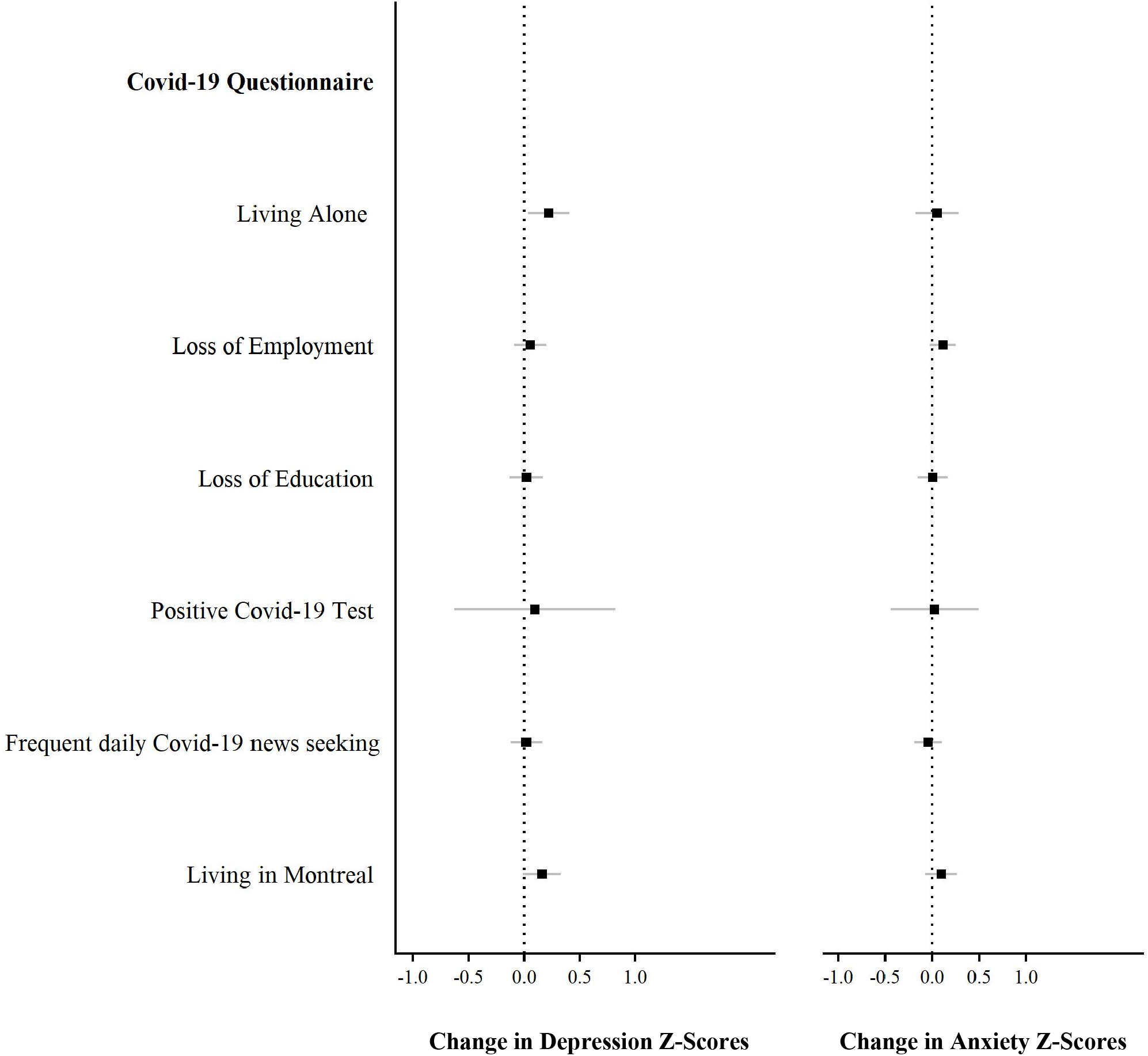
Change in depression and anxiety Z-scores from before (age 20 years) to during the COVID-19 pandemic (age 22 years) as a function of variables in the COVID-19 questionnaire; *n*=1039, weighted. *Note*: Data were compiled from the final master file of the Québec Longitudinal Study of Child Development (1998–2020), Québec Government, Québec Statistic Institute. Positive scores indicate a deterioration of mental health.

**Figure 3.**
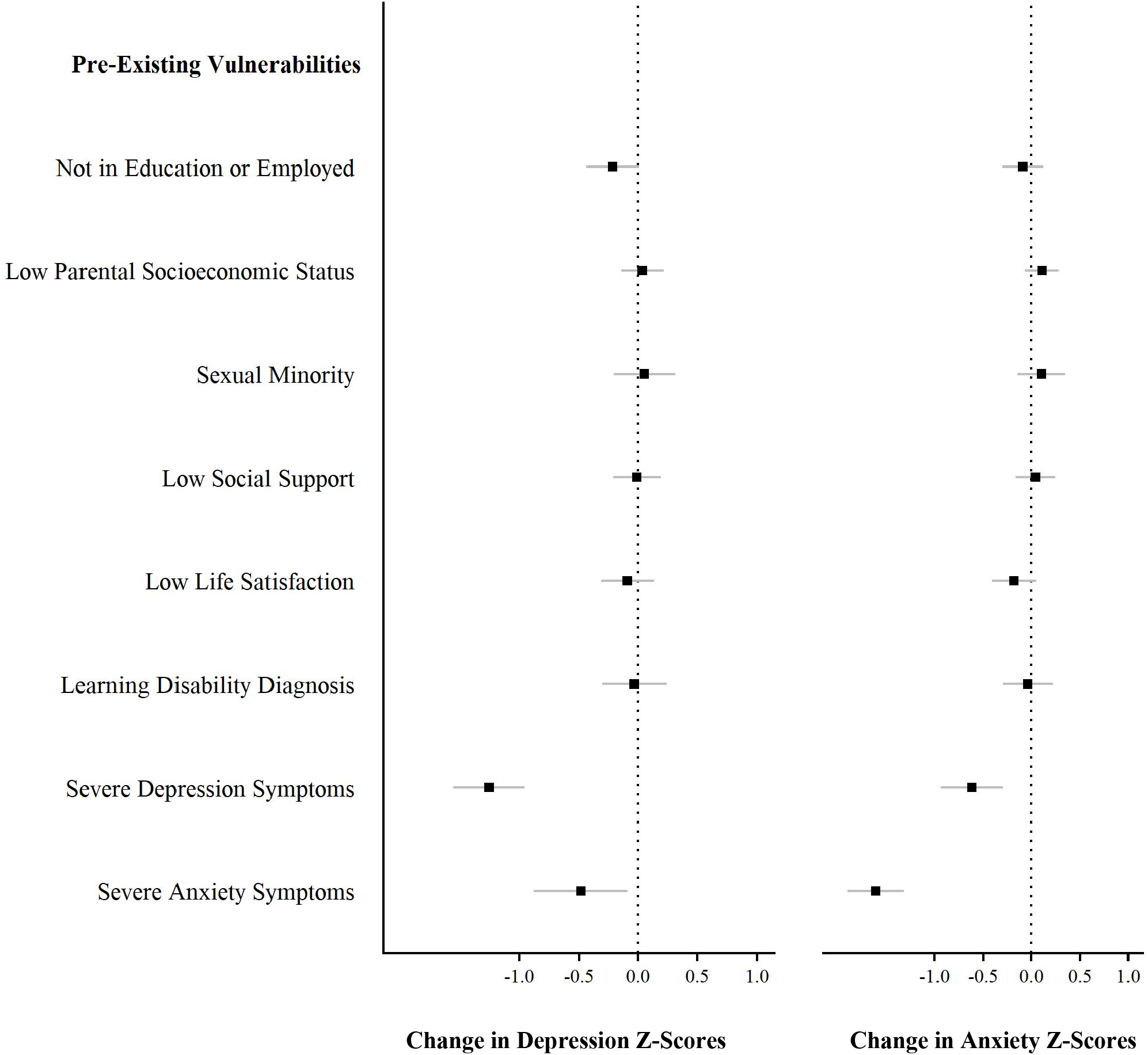
Change in depression and anxiety Z-scores from before (age 20 years) to during the COVID-19 pandemic (age 22 years) as a function of pre-existing vulnerabilities; *n*=1039, weighted. *Note*: Data were compiled from the final master file of the Québec Longitudinal Study of Child Development (1998–2020), Québec Government, Québec Statistic Institute. Positive scores indicate a deterioration of mental health.

**Figure 4.**
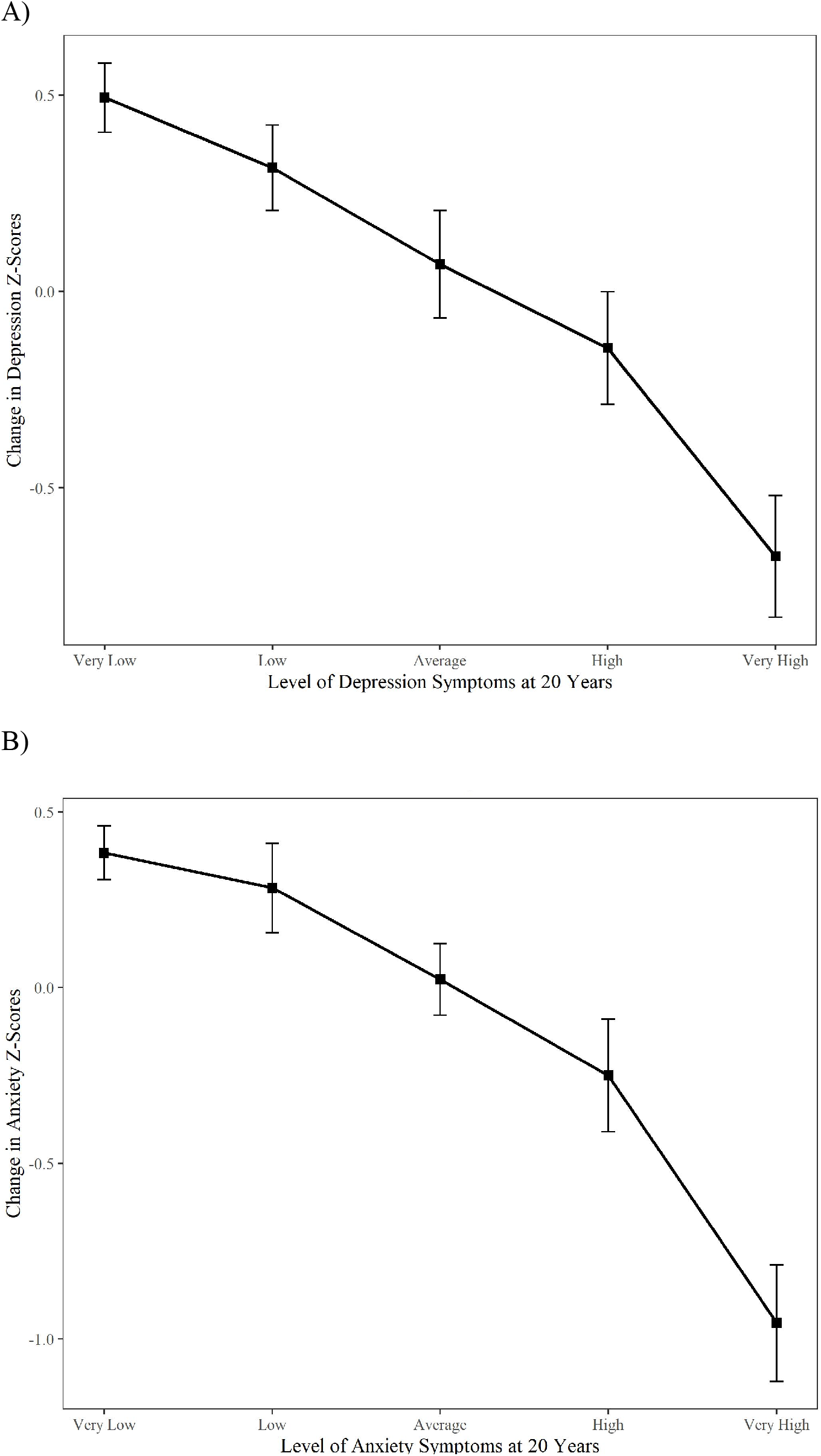
Change in (A) depression and (B) anxiety Z-scores from before (age 20 years) to during the COVID-19 pandemic (age 22 years) as a function of categories of levels of pre-existing symptoms; *n*=1039, weighted. *Note*: Data were compiled from the final master file of the Québec Longitudinal Study of Child Development (1998–2020), Québec Government, Québec Statistic Institute. Positive scores indicate a deterioration of mental health.

## Discussion

Using a population-based cohort of young adults with data collected two years before the pandemic and 4 months after the onset of the first wave of the pandemic (Summer 2020) in the province of Quebec, this study examined changes in symptoms of depression and anxiety and investigated whether these changes are linked to pandemic-related stressors and/or pre-existing vulnerabilities. On average, young adults did not report change in depression and anxiety levels across the full spectrum of symptom severity. However, the prevalence of severe depressive symptoms increased by 1.9% during the initial months of the pandemic, while there was no change in severe anxiety symptoms. Most COVID-19 related stressors (e.g., loss of education/occupation) and variables pertaining to pre-existing vulnerabilities (e.g., low SES, low social support) were not associated with changes in symptoms of depression and anxiety. Contrary to expectations (Copeland et al., 2021; Shanahan et al., 2020), we did not observe a generalized worsening of depressive and anxiety symptoms during the pandemic among participants with severe pre-pandemic depression and anxiety. However, supplemental analyses investigating changes in depression and anxiety symptoms according to different levels of pre-existing symptoms (very low; low; average; high; very high) revealed that young adults with low levels of symptoms before the pandemic experienced a deterioration in mental health during the pandemic. Conversely, those with high levels of pre-existing symptoms at age 20 years experienced an improvement at age 22 years.

Our findings showing no increase in depression and anxiety symptoms from before to during the pandemic are comparable to a prior longitudinal study (Shanahan et al., 2020) using a sample of 768 young adults living in Switzerland. This study found that internalizing symptoms (assessed using depressive, anxiety, and suicidal ideation and self-injury items from the Social Behavior Questionnaire) did not significantly increase from before (20 years) to during the pandemic in April 2020 (22 years). In contrast, a study of 624 undergraduate students (mean age: 19.6 years) in Baoding, China found that symptoms of depression and anxiety (as measured by the PHQ-4) significantly increased from before (December 2019) to during the pandemic (February 2020) (Li et al., 2020).

Although on average we did not observe changes across the full spectrum of symptom severity, the prevalence of severe depression slightly increased by 1.9% from before to during the pandemic. However, the prevalence of severe anxiety did not change, suggesting that mental health consequences of the pandemic might differ depending on the type of symptom. This is in line with a recent systematic review and meta-analysis of 65 longitudinal cohort studies by Robinson and colleagues that found that compared to symptoms of anxiety, increases in depressive symptoms tended to be larger and remained elevated beyond the first months of the pandemic (Robinson et al., 2021).

None of the Covid-19 related stressors were associated with a change in depression and anxiety symptoms, except for those living alone who experienced a slight increase in depressive symptoms. These findings are in line with results from a large prospective panel study weighted to population proportions (n=36,520) and conducted over the first 20 weeks of lockdown in the UK that identified living alone as a risk factor for higher symptoms of depression and anxiety during the pandemic. While most pre-existing vulnerabilities were not associated with change in depression and anxiety symptoms, we found that participants with pre-existing severe manifestations of depression and anxiety at age 20 years experienced a significant decrease in depressive and anxiety symptoms between 2018 and the summer of 2020. Pre-existing mental health vulnerabilities have been identified in several cross-sectional studies as a risk factor for mental health deterioration during the pandemic (González-Sanguino et al., 2020; Özdin and Bayrak Özdin, 2020; Robillard et al., 2021; Varma et al., 2021; Zhou et al., 2020), yet evidence from longitudinal studies is relatively scarce and contradictory, to date (Robinson et al., 2021). For example, in a longitudinal study conducted on three psychiatry case control cohorts in the Netherlands (*N*=1517, mean age: 56.1 years), the COVID-19 pandemic did not seem to exacerbate the severity of pre-existing symptoms among those with severe pre-pandemic mental health problems (Pan et al., 2021), which researchers suggested might be explained by stay-at-home orders allowing for the implementation of more structured and consistent daily routines (Pan et al., 2021; Pelto-Piri et al., 2019). In contrast, a cohort study on young adults in Switzerland identified pre-pandemic emotional distress as the largest risk factor for increased emotional distress during the pandemic (Shanahan et al., 2020).

In our study, symptoms of youth with severe pre-existing symptoms did not deteriorate but rather improved, possibly reflecting a natural improvement of mental illness over time (regression to the mean), an effect of seasonality (2018 data was collected in the spring, while 2020 data was collected in the summer), or a reduction in social stressors during periods of confinement. Conversely, young adults with low levels of depression and anxiety symptoms experienced a significantly greater deterioration in mental health from before to during the pandemic. These results correspond with findings from a longitudinal study by Hamza and colleagues conducted with students at a Canadian university (*N*=773), in which participants with pre-existing mental health problems showed decreasing depressive and anxiety symptoms (from before (May 2019) to during (May 2020) the pandemic, whereas their peers without pre-existing mental health problems showed increasing depressive and anxiety symptoms during the pandemic (Hamza et al., 2020). These findings might reflect the negative impact of increased social isolation (due to widespread closures and social distancing regulations) on young adults without pre-existing mental health problems.

Altogether, these conflicting findings about the mental health consequences of the pandemic may be attributed to several different factors. Firstly, changes in depression and anxiety symptoms might vary according to the type and severity of government lockdown measures in place when symptoms were assessed (Benke et al., 2020). Indeed, the systematic review of longitudinal studies by Robinson and colleagues mentioned above found that while there was an overall increase in mental health symptoms from before to during pandemic, this increase was most pronounced during the early stages of the pandemic (March-April 2020) before decreasing back toward pre-pandemic levels over the following months (May-July 2020). Data for the current study was collected in July-August 2020, during which time Quebecers were newly permitted to gather in private (up to 10 people from 3 different households) and public settings (up to 50 people in movie theaters, reception halls, and places of worship, among other locations) after months of strict lockdown (“Liste de toutes les infographies en lien avec les annonces du premier ministre (COVID-19),” 2021). Participants’ newfound ability to socialize after months of isolation may have offset the impact of pandemic-related stress experienced during the strict lockdown period. Indeed, a German study examining the effects of different kinds and levels of restrictive public health measures during the pandemic (e.g., quarantine/lockdown/stay-at-home orders) on symptoms of anxiety and depression found that stricter restrictions, greater reduction of social contact, and greater perceived changes in daily life were associated with poorer mental health (Benke et al., 2020).

Secondly, depression and anxiety symptoms may vary in accordance with government socioeconomic policies, which differ across countries. Soon after the onset of the pandemic, the Canadian government introduced relatively generous payments for workers and post-secondary students whose income earning abilities were negatively impacted (e.g., reduction in hours; job loss) by the COVID-19 pandemic (Canada Revenue Agency, 2020; Government of Canada, 2020). These government measures may have had a protective effect against depression and anxiety. For example, a survey study using data collected by the US census Bureau from April to July 2020 found that the prevalence of depressive and anxiety symptoms varied across states by what the investigators called “household income shock” (Donnelly and Farina, 2021) (i.e. the experience of job loss and/or partial income loss), which was buffered by state-level differing socioeconomic policies (e.g. access to Medicaid, unemployment insurance, and suspended utility shut-offs) (Donnelly and Farina, 2021). Despite the potentially buffering effects of the Canadian government’s economic response plan against symptoms of depression and anxiety, it is worth noting that future professional opportunities were still among the primary COVID-related concerns shared by young adults, with 21.4% of our participants stating that they were very or extremely concerned by future job prospects (see **Figure 1**).

Lastly, the normal process of psychological adaptation following distressing events might also come into play in explaining the inconsistent findings on the impact of the pandemic on the mental health of young adults (Daly and Robinson, 2021; Pierce et al., 2020). Consistent with findings from a meta-analysis indicating that mental health deteriorated at the onset of the pandemic before returning to baseline levels by mid-2020 (Robinson et al., 2021), a prospective longitudinal study in the UK examining anxiety and depression over the first 20 weeks of lockdown (March 23-August 9, 2020) found that there was a significant decrease in depressive and anxiety symptoms throughout both the strict lockdown period and the period during which lockdown measures were eased (Fancourt et al., 2021). Moreover, the same study found that younger adults showed faster improvements in depression and anxiety symptoms compared to older adults (Fancourt et al., 2021). This suggests that while some studies find young adults to be at greater risk for mental health problems during the pandemic, they may be better able to psychologically adapt to challenging circumstances relative to older adults, or that factors contributing to persistent internalizing symptoms may have been alleviated by the confinement measures (e.g., lack of commute; additional rest time; accommodations for final exams). However, given that most studies on the mental health impact of the COVID-19 pandemic on young adults were conducted during the first wave of the pandemic in 2020, the mental health impact of the subsequent waves is still unclear.

### Methodological Considerations

This study has a number of strengths, including its longitudinal design; the use of standardized measures of depression and anxiety; and data collected before and during the pandemic from the same participants. We acknowledge the following limitations. As in all longitudinal surveys, attrition occurred over the years and the most vulnerable individuals were underrepresented. Although all analyses were weighted, such differential attrition could potentially result in underestimation of the rates of anxiety and depression and consequently of the mental impact of COVID-19. Mental health outcomes were measured by self-report questionnaires, which do not provide clinical diagnoses. While we were able to use longitudinal data with mental health assessments before and during the Covid-19 pandemic, it is difficult to differentiate between mental health changes attributable to the pandemic versus developmental changes that are typical for this age group or changes due to seasonal effects, since pre-pandemic measures were obtained in the Spring, while intra-pandemic measures were collected during the Summer.

### Conclusion

On average, depressive and anxiety symptoms did not significantly change during the first wave of the Covid-19 pandemic among young adults, although the prevalence of severe depression did increase. Moreover, while the majority of the young adults showed a pattern of symptoms consistent with adaptation to the pandemic, levels of depressive and anxiety symptoms increased among those with the lowest levels of symptoms before the pandemic. These findings underscore the need for facilitating access to treatment for the subgroup of individuals who may be newly struggling with symptoms of depression and anxiety. Future studies should track the mental health of young adults throughout the subsequent waves of the COVID-19 pandemic beyond the summer of 2020.

## Data Availability

Data are managed by Institut de la statistique du Quebec (ISQ) and data are accessible upon approval by the ISQ.

## Acknowledgements

The larger Quebec Longitudinal Study of Child Development was supported by funding from the Quebec provincial government’s Ministry of Health and Ministry of Family Affairs, the Lucie and André Chagnon Foundation, the Fonds de Recherche du Québec en Société et Culture, Canada’s Social Sciences and Humanities Research Council, the Canadian Institutes of Health Research, the Centre Hospitalier Universitaire de Sainte-Justine, and the Institut de la Statistique du Québec. The authors thank Alain Girard and Elise Chartrand for their contributions to the statistical analysis and all participants of the Quebec Longitudinal Study of Child Development.

---

**Supplemental Table 1.** Change in depression and anxiety symptoms from before to during the COVID-19 pandemic.

| Variables | Mean difference:<br>depressive<br>symptoms (SE) | *p* value | Mean difference:<br>anxiety symptoms<br>(SE) | *p* value |
| --- | --- | --- | --- | --- |
| **COVID-19 questionnaire** |  |  |  |  |
| Living alone | 0.22 (0.10) | .020 | 0.05 (0.12) | .657 |
| Loss of employment | 0.06 (0.07) | .451 | 0.12 (0.07) | .107 |
| Loss of education | 0.02 (0.08) | .769 | 0.00 (0.08) | .959 |
| Positive COVID-19 test | 0.10 (0.37) | .795 | 0.03 (0.24) | .917 |
| Frequent Daily COVID-related<br>news seeking | 0.02 (0.07) | .772 | -0.04 (0.07) | .560 |
| Living in Montreal | 0.16 (0.09) | .063 | 0.10 (0.09) | .262 |
| **Pre-existing vulnerabilities** |  |  |  |  |
| Not in education or employed | -0.22 (0.12) | .060 | -0.09 (0.11) | .398 |
| Low family SES | 0.04 (0.09) | .686 | 0.10 (0.09) | .239 |
| Sexual orientation minority | 0.05 (0.13) | .680 | 0.10 (0.13) | .435 |
| Learning disability diagnosis | -0.03 (0.14) | .824 | -0.04 (0.13) | .770 |
| Low social support | -0.01 (0.10) | .924 | 0.04 (0.10) | .712 |
| Low life satisfaction | -0.09 (0.11) | .428 | -0.18 (0.12) | .118 |
| Severe pre-existing depression | -1.26(0.15) | <.001 | -0.62 (0.16) | <.001 |
| Severe pre-existing anxiety | -0.48 (0.20) | .016 | -1.61 (0.15) | <.001 |

*Note:* Data were compiled from the final master file of the Québec Longitudinal Study of Child Development (1998–2020), Québec Government, Québec Statistic Institute. Positive scores indicate a deterioration of mental health.

**Supplemental Table 2.** Change in depression and anxiety symptoms from before the pandemic to during the pandemic according to different levels of symptoms severity before the pandemic.

| Symptom Severity Before the Pandemic | Mean difference:<br>depressive symptoms<br>(SE) | *p* value | Mean difference:<br>anxiety symptoms<br>(SE) | *p* value |
| --- | --- | --- | --- | --- |
| Very Low (0-20%) | 0.49 (0.05) | <.001 | 0.38 (0.04) | <.001 |
| Low (21-40%) | 0.32 (0.06) | <.001 | 0.28 (0.06) | <.001 |
| Average (41-60%) | 0.07 (0.07) | .322 | 0.02 (0.05) | .654 |
| High (61-80%) | -0.14 (0.07) | .046 | -0.25 (0.08) | .002 |
| Very High (81-100%) | -0.68 (0.08) | <.001 | -0.96 (0.08) | <.001 |

*Note:* Data were compiled from the final master file of the Québec Longitudinal Study of Child Development (1998–2020), Québec Government, Québec Statistic Institute. Positive scores indicate a deterioration of mental health.

